# Neurological Manifestations of Hospitalized Patients with COVID-19 in Wuhan, China: a retrospective case series study

**DOI:** 10.1101/2020.02.22.20026500

**Authors:** Ling Mao, Mengdie Wang, Shengcai Chen, Quanwei He, Jiang Chang, Candong Hong, Yifan Zhou, David Wang, Yanan Li, Huijuan Jin, Bo Hu

**Affiliations:** Department of neurology, Union hospital, Tongji medical college, Huazhong university of science and technology, Wuhan, 430022, China; Department of Epidemiology and Biostatistics, Key Laboratory for Environment and Health, School of Public Health, Tongji medical college, Huazhong university of science and technology, Wuhan, 430022, China; Neurovascular Division, Department of Neurology, Barrow Neurological Institute/Saint Joseph Hospital Medical Center Phoenix, AZ 85013 USA

## Abstract

**OBJECTIVE:** To study the neurological manifestations of patients with coronavirus disease 2019 (COVID-19).

**DESIGN:** Retrospective case series

**SETTING:** Three designated COVID-19 care hospitals of the Union Hospital of Huazhong University of Science and Technology in Wuhan, China.

**PARTICIPANTS:** Two hundred fourteen hospitalized patients with laboratory confirmed diagnosis of severe acute respiratory syndrome from coronavirus 2 (SARS-CoV-2) infection. Data were collected from 16 January 2020 to 19 February 2020.

**MAIN OUTCOME MEASURES:** Clinical data were extracted from electronic medical records and reviewed by a trained team of physicians. Neurological symptoms fall into three categories: central nervous system (CNS) symptoms or diseases (headache, dizziness, impaired consciousness, ataxia, acute cerebrovascular disease, and epilepsy), peripheral nervous system (PNS) symptoms (hypogeusia, hyposmia, hypopsia, and neuralgia), and skeletal muscular symptoms. Data of all neurological symptoms were checked by two trained neurologists.

**RESULTS:** Of 214 patients studied, 88 (41.1%) were severe and 126 (58.9%) were non-severe patients. Compared with non-severe patients, severe patients were older (58.7 ± 15.0 years vs 48.9 ± 14.7 years), had more underlying disorders (42 [47.7%] vs 41 [32.5%]), especially hypertension (32 [36.4%] vs 19 [15.1%]), and showed less typical symptoms such as fever (40 [45.5%] vs 92 [73%]) and cough (30 [34.1%] vs 77 [61.1%]). Seventy-eight (36.4%) patients had neurologic manifestations. More severe patients were likely to have neurologic symptoms (40 [45.5%] vs 38 [30.2%]), such as acute cerebrovascular diseases (5 [5.7%] vs 1 [0.8%]), impaired consciousness (13 [14.8%] vs 3 [2.4%]) and skeletal muscle injury (17 [19.3%] vs 6 [4.8%]).

**CONCLUSION:** Compared with non-severe patients with COVID-19, severe patients commonly had neurologic symptoms manifested as acute cerebrovascular diseases, consciousness impairment and skeletal muscle symptoms.

## Introduction

In December 2019, many unexplained pneumonia cases occurred in Wuhan, China, and has rapidly spread to other parts of China, then to Europe, North America and Asia. This outbreak was confirmed to be caused by a novel coronavirus (2019 novel coronavirus, 2019-nCoV) [1]. 2019-nCov was reported to have symptoms resembled that of severe acute respiratory syndrome coronavirus (SARS-CoV) in 2003 [2]. Both shared the same receptor, angiotensin-converting enzyme 2 (ACE2) [3]. Therefore, this virus was named SARS-CoV-2, and recently WHO named it coronavirus disease 2019 (COVID-19). Until February 21^th^ 2020, there were 75569 confirmed cases of COVID-19 and 2239 deaths in China [4].

Coronaviruses can cause multiple systemic infections or injuries in various animals [5]. However, some of them can adapt fast and cross the species barrier, such as in the cases of SARS-CoV and Middle East respiratory syndrome-CoV (MERS-CoV), causing epidemics or pandemics. Infection in human often leads to severe clinical symptoms and high mortality [6]. As for COVID-19, several studies have described clinical manifestations including respiratory symptoms, myalgia and fatigue. COVID-19 also has characteristic laboratory findings and lung CT abnormalities [7]. However, it has not been reported that patients with COVID-19 had any neurological manifestations. Here, we would like to report the characteristic neurological manifestation of SARS-CoV-2 infection in 78 of 214 patients with laboratory-confirmed diagnosis of COVID-19 and treated at our hospitals, which are located in the epicenter of Wuhan.

## Methods

### Study Design and Participants

This was a retrospective study. Data was reviewed on all patients with COVID-19 from January 16 to February 19, 2020 at three designated COVID-19 care hospitals of Union Hospital of Huazhong University of Science and Technology. All patients with COVID-19 enrolled in this study were diagnosed according to the WHO interim guideline [8]. Only those cases confirmed by a positive result to real-time reverse-transcriptase polymerase-chain-reaction (RT-PCR) assay from throat swab specimens were included in the analysis [9]. Union Hospital, located in the endemic areas of COVID-19 in Wuhan, Hubei Province, is one of the major tertiary healthcare system and teaching hospitals responsible for the treatments for SARS-CoV-2 infection as designated by the government. The study was performed in accordance to the principles of the Declaration of Helsinki and was approved by the Research Ethics Committee of Tongji Medical College, Huazhong University of Science and Technology, Wuhan, China. Verbal consent was obtained from patients before the enrollment.

### Data Collection

The demographic characteristics, medical history, symptoms, clinical signs, laboratory findings, chest computed tomographic (CT) scan findings were extracted from electronic medical records. The data were reviewed by a trained team of physicians. Neurological symptoms were categorized into three main areas: central nervous system (CNS) symptoms or disease, peripheral nervous system (PNS) symptoms and muscular symptoms. Acute cerebrovascular disease included ischemic stroke and cerebral hemorrhage diagnosed by head CT. Muscle injury was defined when a patient had myalgia and elevated serum creatine kinase level above 200 U/L [7]. All neurological symptoms were reviewed and confirmed by two trained neurologists. The date of disease onset was defined as the day when the symptom was noticed. The severity of COVID-19 was defined by the international guidelines for community-acquired pneumonia [10].

Throat swab samples were collected and placed into a collection tube containing preservation solution for the virus [9]. SARS-CoV-2 was confirmed by real-time RT-PCR assay using a SARS-CoV-2 nucleic acid detection kit according to the manufacturer’s protocol (Shanghai bio-germ Medical Technology Co Ltd).

### Statistical Analysis

Continuous variables were described as means and standard deviations, or medians and interquartile range (IQR) values. Categorical variables were expressed as counts and percentages. Continuous variables were compared by using the unpaired Wilcox rank-sum test. Proportions for categorical variables were compared using the χ2 test. All statistical analyses were performed using R (version 3.3.0) software. The significance threshold was set at a *P*<0.05.

## Results

### Demographic and clinical characteristics

A total of 214 hospitalized patients with confirmed SARS-CoV-2 infection were included in the present analysis. Their demographic and clinical characteristics were shown in Table 1. Their average age was 52.7 ± 15.5 years, and 127 (59.3%) were females. Of these patients, 83 (38.8%) had at least one of the following underlying disorders: hypertension (51 [23.8%]), diabetes (30 [14.0%]), cardiovascular disease (15 [7.0%]), and malignancy (13 [6.1%]). The most common symptoms at onset of illness were fever (132 [61.7%]), dry cough (107 [50.0%]) and anorexia (68 [31.8%]). Seventy-eight (36.4%) patients had nervons system symptoms: CNS (53 [24.8%]), PNS (19 [8.9%]) and skeletal muscles (23 [10.7%]). In patients with CNS symptoms, the most common complaints were dizziness (36 [16.8%] and headache (28 [13.1%]). In patients with PNS symptoms, the most common complaints were hypogeusia (12 [5.6%]) and hyposmia (11 [5.1%]).

**Table 1.**
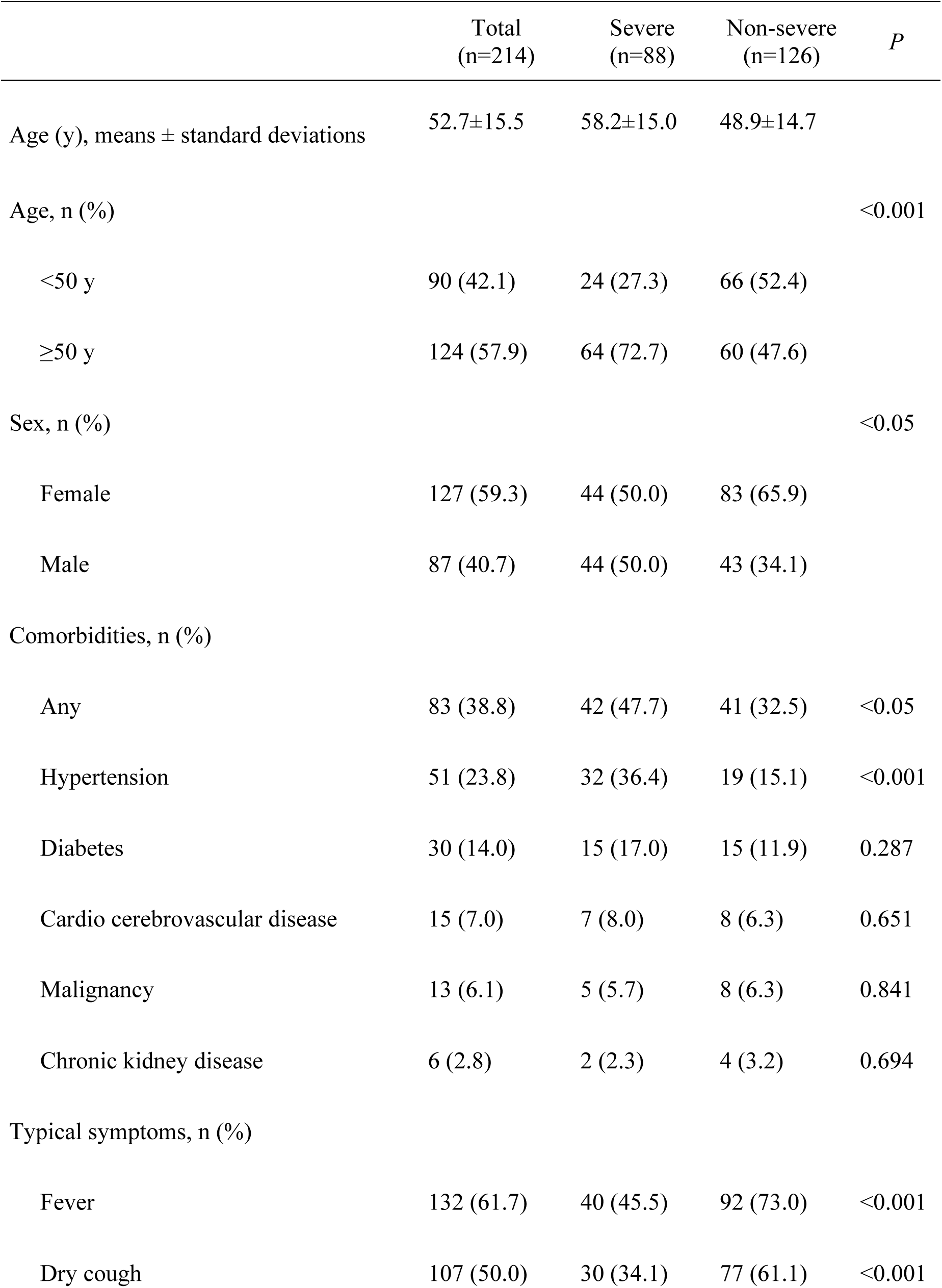
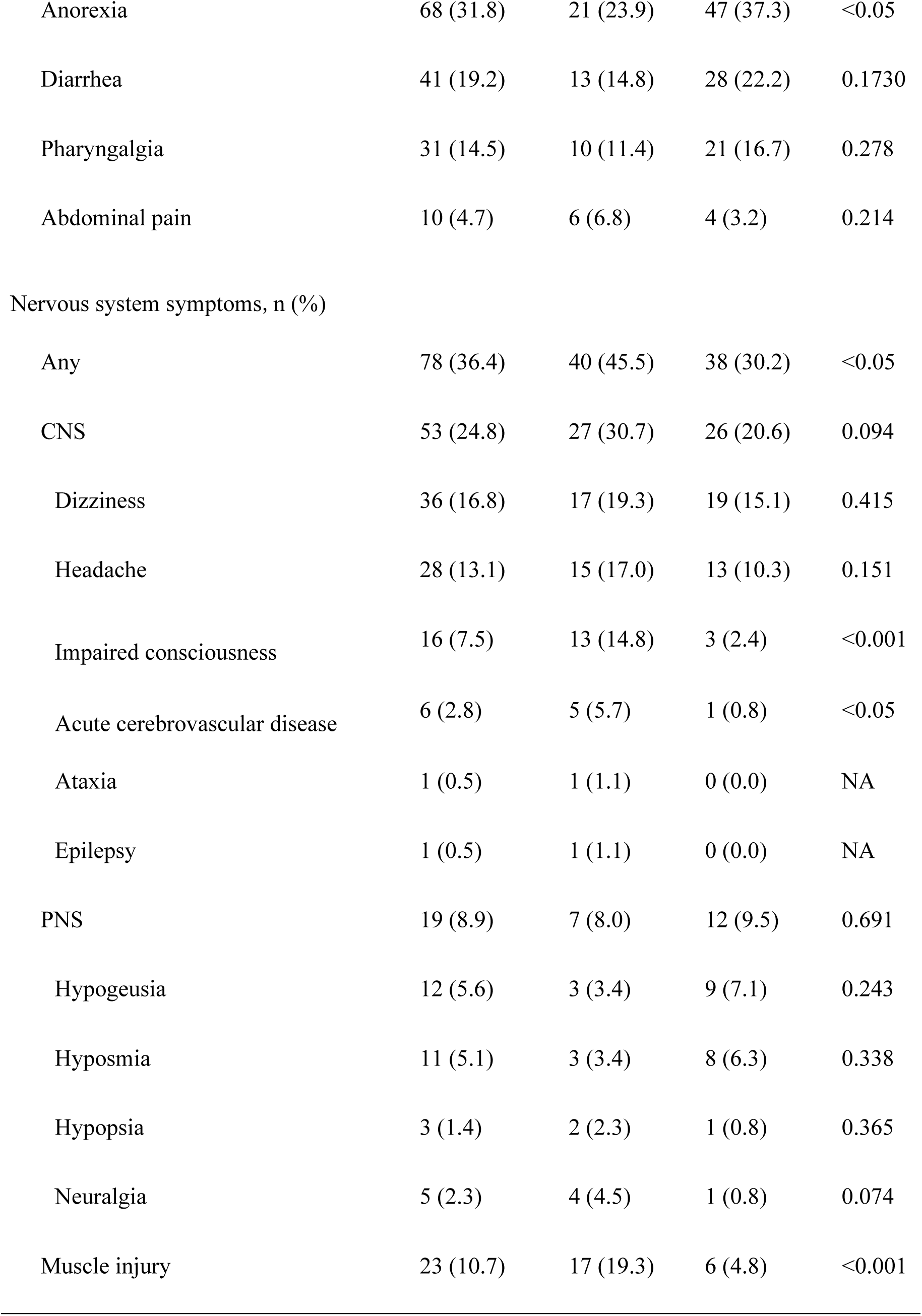
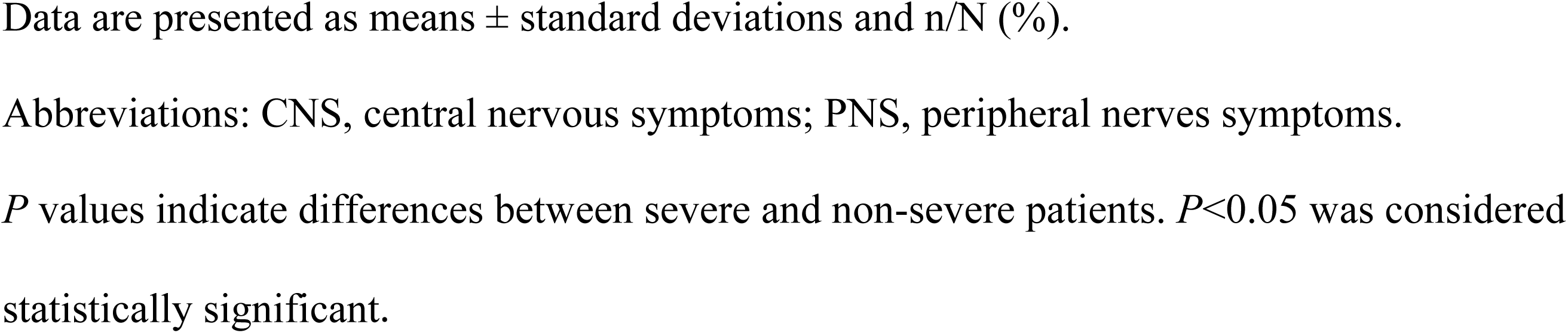
Clinical characteristics of patients with COVID-19

According to the diagnostic criteria, 88 (41.1%) patients were severe and 126 (58.9%) patients were non-severe, respectively. The patients with severe infection were significantly older (58.2 ± 15.0 years vs 48.9 ± 14.7 years; *P*<0.001) and more likely to have other underlying disorders (42 [47.7%] vs 41 [32.5%], *P*<0.05), especially hypertension (32 [36.4%] vs 19 [15.1%], *P*<0.001), and had less typical symptoms such as fever (40 [45.5%] vs 92 [73%], *P*<0.001) and dry cough (30 [34.1%] vs 77 [61.1%], *P*<0.001). Moreover, nervous system symptoms were significantly more common in severe cases as compared with non-severe cases (40 [45.5%] vs. 38 [30.2%], *P*<0.05). They included acute cerebrovascular disease (5 [5.7%] (4 patients with ischemic stroke and 1 with cerebral hemorrhage who died later from respiratory failure) vs. 1 [0.8%] (1 patient with ischemic stroke), *P*<0.05), impaired consciousness (13 [14.8%] vs. 3 [2.4%], *P*<0.001) and muscle injury (17 [19.3%] vs. 6 [4.8%], *P*<0.001).

### Laboratory findings in severe patients and non-severe patients

Table 2 showed the laboratory findings in severe and non-severe subgroups. Severe patients had more increased inflammatory response, including higher white blood cell, neutrophil counts, lower lymphocyte counts and more increased C-reaction protein levels compared with those in non-severe patients (white blood cell: median, 5.4 [IQR, 0.1-20.4] vs 4.5 [IQR, 1.8-14.0], *P*<0.01; neutrophil: median, 3.8 [IQR, 0.0-18.7] vs 2.6 [IQR, 0.7-11.8], *P*<0.001; lymphocyte: median, 0.9 [IQR, 0.1-2.6] vs 1.3 [IQR, 0.4-2.6], *P*<0.001; C-reaction protein: median, 37.1 [IQR, 0.1-212.0] vs 9.4 [IQR, 0.2-126.0], *P*<0.001). The severe patients had higher D-dimer levels than non-severe patients (median, 0.9 [IQR, 0.1-20.0] vs 0.4 [IQR, 0.2-8.7], *P*<0.001), which was indicative of consumptive coagulation system. In addition, severe patients had multiple organ involvement, such as serious liver (increased lactate dehydrogenase, alanine aminotransferase and aspartate aminotransferase levels), kidney (increased blood urea nitrogen and creatinine levels) and muscle damage (increased creatinine kinase levels).

**Table 2.** Laboratory findings of patients with COVID-19

| Median (range) | Total<br>(n=214) | Severe<br>(n=88) | Non-severe<br>(n=126) | *P* |
| --- | --- | --- | --- | --- |
| White blood cell count,<br>×10<sup>9</sup>/L | 4.9 (0.1-20.4) | 5.4 (0.1-20.4) | 4.5 (1.8-14.0) | <0.01 |
| Neutrophil, ×10<sup>9</sup>/L | 3.0 (0.0-18.7) | 3.8 (0.0-18.7) | 2.6 (0.7-11.8) | <0.001 |
| Lymphocyte count, ×10<sup>9</sup>/L | 1.1 (0.1-2.6) | 0.9 (0.1-2.6) | 1.3 (0.4-2.6) | <0.001 |
| Platelet count, ×10<sup>9</sup>/L | 209.0 (18.0-583.0) | 204.5 (18.0-576.0) | 219.0 (42.0-583.0) | 0.251 |
| C-reactive protein, mg/L | 12.2 (0.1-212.0) | 37.1 (0.1-212.0) | 9.4 (0.4-126.0) | <0.001 |
| D-dimer, mg/L | 0.5 (0.1-20.0) | 0.9 (0.1-20.0) | 0.4 (0.2-8.7) | <0.001 |
| Creatine kinase, U/L | 64.0 (8.8-12216.0) | 83.0 (8.8-12216.0) | 59.0 (19.0-1260.0) | <0.01 |
| Lactate dehydrogenase,<br>U/L | 241.5 (2.2-908.0) | 302.0 (2.2-880.0) | 215.0 (2.5-908.0) | <0.001 |
| Alanine aminotransferase,<br>U/L | 26.0 (5.0-1933.0) | 32.5 (5.0-1933.0) | 23.0 (6.0-261.0) | <0.05 |
| Aspartate aminotransfer-<br>ase, U/L | 26.0 (8.0-8191.0) | 34.0 (8.0-8191.0) | 23.0 (9.0-244.0) | <0.001 |
| Blood urea nitrogen,<br>mmol/L | 4.1 (1.5-48.1) | 4.6 (1.5-48.1) | 3.8 (1.6-13.7) | <0.001 |
| Creatinine, $\mu\text{mol/L}$ | 68.2 (35.9-9435.0) | 71.6 (35.9-9435.0) | 65.6 (39.4-229.1) | <0.05 |

*P* values indicate differences between severe and non-severe patients. $P<0.05$ was considered statistically significant.

### Laboratory findings in patients with and without CNS symptoms

Table 3 showed the laboratory findings of patients with and without CNS symptoms. We found that patients with CNS symptoms had lower lymphocyte, platelet counts and higher blood urea nitrogen levels compared with those without CNS symptoms (lymphocyte: median, 1.0 [IQR, 0.1-2.3] vs 1.2 [IQR, 0.2-2.6], *P*<0.05; platelet: median, 180.0 [IQR, 18.0-564.0] vs 227.0 [IQR, 42.0-583.0], *P*<0.01; blood urea nitrogen: median, 4.5 [IQR, 1.6-48.1] vs 4.1 [IQR, 1.5-19.1], *P*<0.05). For the severe sub-group, patients with CNS symptoms also had lower lymphocyte, platelet counts and higher blood urea nitrogen levels compared with those without CNS symptoms (lymphocyte: median, 0.7 [IQR, 0.1-1.6] vs 0.9 [IQR, 0.2-2.6], *P*<0.01; platelet: median, 169.0 [IQR, 18.0-564.0] vs 220.0 [IQR, 109.0-576.0], *P*<0.05; blood urea nitrogen: median, 5.0 [IQR, 2.3-48.1] vs 4.4 [IQR, 1.5-19.1], *P*<0.05). For non-severe subgroup, there were no significant differences in laboratory findings of patients with and without CNS symptoms.

**Table 3.** Laboratory findings of COVID-19 patients with CNS symptoms

|  | Total |  |  | Severe |  |  | Non-severe |  |  |
| --- | --- | --- | --- | --- | --- | --- | --- | --- | --- |
|  | With CNS symptoms (n=53) | Without CNS symptoms (n=161) | *P* | With CNS symptoms (n=27) | Without CNS symptoms (n=61) | *P* | With CNS symptoms (n=26) | Without CNS symptoms (n=100) | *P* |
| Median (range) |  |  |  |  |  |  |  |  |  |
| White blood cell count, ×10<sup>9</sup>/L | 4.6 (0.1-12.5) | 4.9 (1.8-20.4) | 0.582 | 5.3 (0.1-12.5) | 5.5 (1.9-20.4) | 0.769 | 4.1 (2.4-11.0) | 4.6 (1.8-14.0) | 0.397 |
| Neutrophil, ×10<sup>9</sup>/L | 2.6 (0.0-10.9) | 3.1 (0.7-18.7) | 0.413 | 3.8 (0.0-10.9) | 3.6 (0.7-18.7) | 1.000 | 2.2 (0.9-7.4) | 2.8 (0.7-11.8) | 0.106 |
| Lymphocyte count, ×10<sup>9</sup>/L | 1.0 (0.1-2.3) | 1.2 (0.2-2.6) | <0.05 | 0.7 (0.1-1.6) | 0.9 (0.2-2.6) | <0.01 | 1.3 (0.7-2.3) | 1.3 (0.4-2.6) | 0.492 |
| Platelet count, ×10<sup>9</sup>/L | 180.0 (18.0-564.0) | 227.0 (42.0-583.0) | <0.01 | 169.0 (18.0-564.0) | 220.0 (109.0-576.0) | <0.05 | 188.5 (110.0-548.0) | 232.0 (42.0-583.0) | 0.093 |
| C-reactive protein, mg/L | 14.1 (0.1-212.0) | 11.4 (0.1-204.5) | 0.307 | 48.6 (0.1-212.0) | 26.1 (0.1-204.5) | 0.677 | 7.4 (3.1-111.0) | 9.8 (0.4-126.0) | 0.817 |
| D-dimer, mg/L | 0.5 (0.2-9.7) | 0.5 (0.1-20.0) | 0.750 | 1.2 (0.2-9.7) | 0.9 (0.1-20.0) | 0.424 | 0.4 (0.2-6.4) | 0.4 (0.2-8.7) | 0.455 |
| Creatine kinase, U/L | 79.0 (8.8-12216.0) | 60.5 (19.0-1260.0) | 0.167 | 104.0 (8.8-12216.0) | 64.0 (19.0-1214.0) | 0.081 | 52.5 (28.0-206.0) | 59.0 (19.0-1260.0) | 0.561 |
| Lactate dehydrogenase, U/L | 243.0 (2.2-880.0) | 241.0 (3.5-908.0) | 0.766 | 334.0 (2.2-880.0) | 299.0 (3.5-743.0) | 0.324 | 198.0 (2.5-417.0) | 226.0 (121.0-908.0) | 0.142 |
| Alanine aminotransferase, U/L | 27.0 (5.0-261.0) | 26.0 (6.0-1933.0) | 0.214 | 35.0 (5.0-259.0) | 31.0 (7.0-1933.0) | 0.320 | 25.5 (13.0-261.0) | 23.0 (6.0-135.0) | 0.682 |
| Aspartate aminotransferase, U/L | 29.5 (13.0-213.0) | 26.0 (8.0-8191.0) | 0.1031 | 35.5 (14.0-213.0) | 34.0 (8.0-8191.0) | 0.319 | 23.0 (13.0-198.0) | 23.5 (9.0-244.0) | 0.575 |
| Blood urea nitrogen, mmol/L | 4.5 (1.6-48.1) | 4.1 (1.5-19.1) | <0.05 | 5.0 (2.3-48.1) | 4.4 (1.5-19.1) | 0.041 | 3.9 (1.6-9.4) | 3.8 (1.7-13.7) | 0.568 |
| Creatinine, μmol/L | 71.7 (37.1-1299.2) | 66.3 (35.9-9435.0) | 0.062 | 71.7 (37.1-1299.2) | 68.4 (35.9-9435.0) | 0.245 | 72.0 (40.3-133.6) | 63.4 (39.4-229.1) | 0.265 |

Abbreviations: CNS, central nerves system.
*P* values indicate differences between patients with and without CNS. *P* < 0.05 was considered statistically significant

### Laboratory findings in patients with and without PNS symptoms

Table 4 showed the laboratory findings of patients with and without PNS symptoms. We found that there were no significant differences in laboratory findings of patients with PNS and those without PNS. Similar results were also found in the severe subgroup and non-severe subgroup, respectively.

**Table 4.**
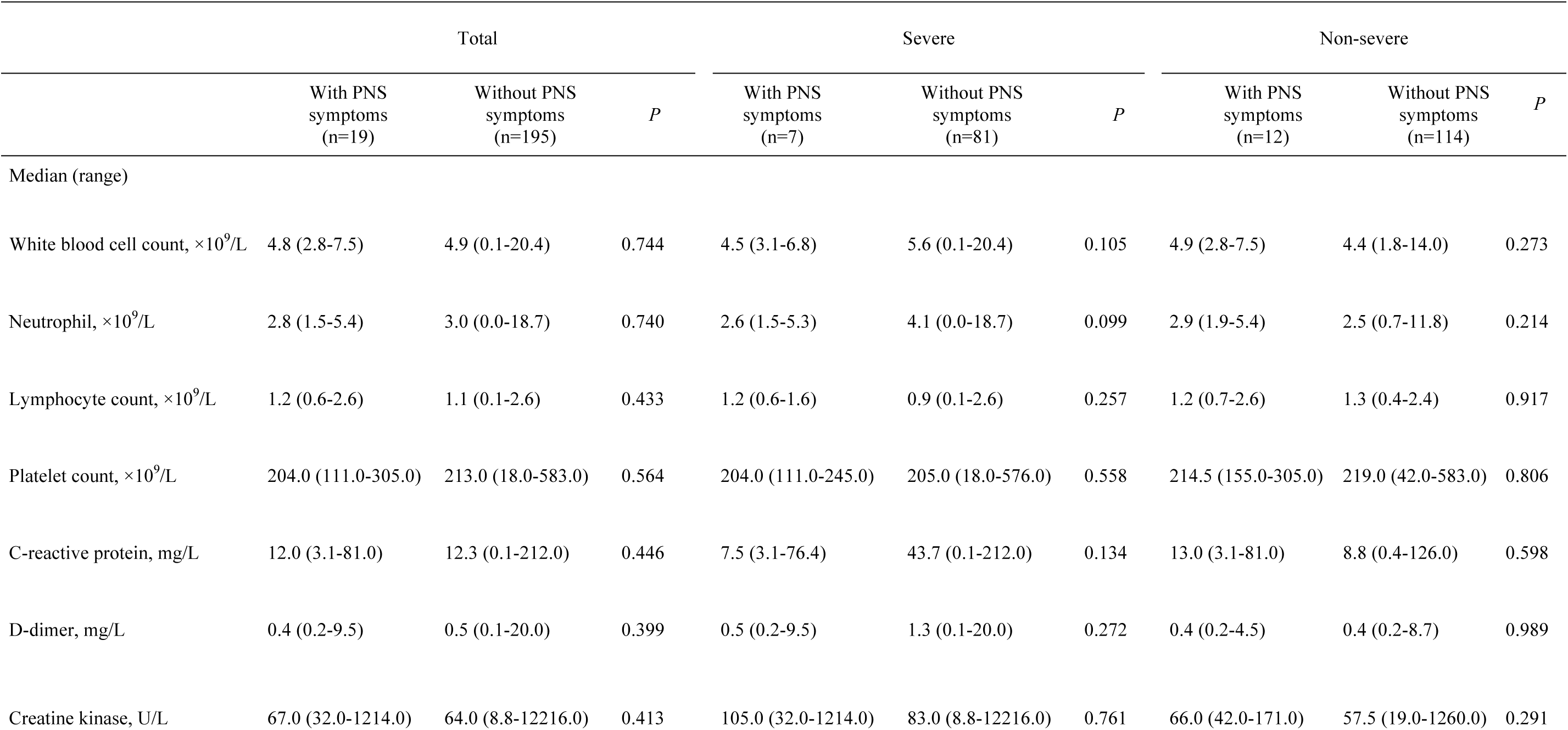
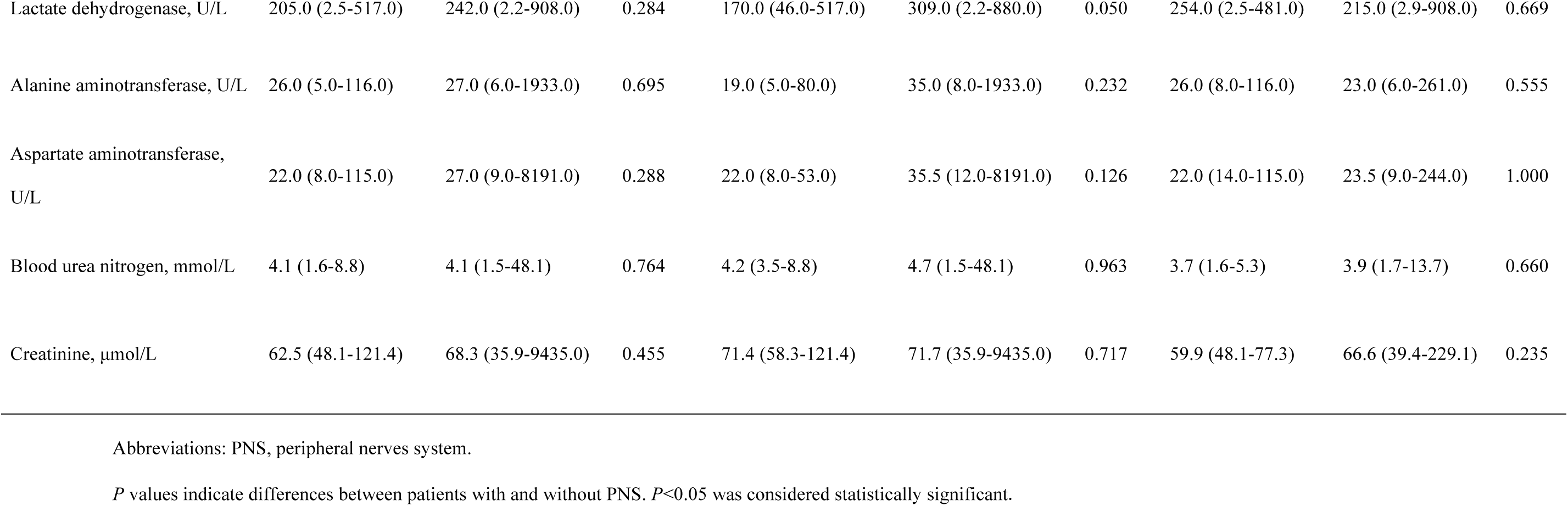
Laboratory findings of COVID-19 patients with PNS symptoms

### Laboratory findings in patients with and without muscle injury

Table 5 showed the laboratory findings of patients with and without muscle injury. Compared with the patients without muscle injury, patients with muscle injury had higher neutrophil counts, lower lymphocyte counts and higher C-reactive protein levels, D-dimer levels (neutrophil: median, 4.3 [IQR, 0.9-18.7] vs 2.9 [IQR, 0.0-13.0], *P*<0.05; lymphocyte: median, 0.9 [IQR, 0.1-2.6] vs 1.2 [IQR, 0.1-2.6], *P*<0.01; C-reaction protein: median, 56.0 [IQR, 0.1-212.0] vs 11.1 [IQR, 0.1-204.5], *P*<0.001; D-dimer: median, 1.3 [IQR, 0.2-20.0] vs 0.5 [IQR, 0.1-20.0]). The abnormalities were manifestation of increased inflammatory response and blood coagulation function. In addition, we found that patients with muscle injury had multi-organ damage including more serious liver (increased lactate dehydrogenase, alanine aminotransferase and aspartate aminotransferase levels), and kidney (increased blood urea nitrogen and creatinine levels) abnormalities.

**Table 5.** Laboratory findings of COVID-19 patients with muscle injury

|  | Total |  |  | Severe |  |  | Non-severe |  |  |
| --- | --- | --- | --- | --- | --- | --- | --- | --- | --- |
|  | With muscle injury<br>(n=23) | Without muscle injury<br>(n=191) | *P* | With muscle injury<br>(n=17) | Without muscle injury<br>(n=71) | *P* | With muscle injury<br>(n=6) | Without muscle injury<br>(n=120) | *P* |
| Median (range) |  |  |  |  |  |  |  |  |  |
| White blood cell count, ×10<sup>9</sup>/L | 6.0 (2.3-20.4) | 4.8 (0.1-15.6) | 0.248 | 6.7 (2.3-20.4) | 5.2 (0.1-15.6) | 0.269 | 4.3 (2.4-6.1) | 4.5 (1.8-14.0) | 0.618 |
| Neutrophil, ×10<sup>9</sup>/L | 4.3 (0.9-18.7) | 2.9 (0.0-13.0) | 0.031 | 5.5 (0.9-18.7) | 3.5 (0.0-13.0) | 0.076 | 2.6 (1.1-4.5) | 2.6 (0.7-11.8) | 0.945 |
| Lymphocyte count, ×10<sup>9</sup>/L | 0.9 (0.1-2.6) | 1.2 (0.1-2.6) | 0.002 | 0.8 (0.1-2.6) | 0.9 (0.1-2.5) | <0.05 | 1.2 (1.0-2.1) | 1.3 (0.4-2.6) | 0.846 |
| Platelet count, ×10<sup>9</sup>/L | 185.0 (82.0-436.0) | 215.0 (18.0-583.0) | 0.224 | 182.0 (82.0-436.0) | 209.0 (18.0-576.0) | 0.407 | 223.0 (122.0-304.0) | 219.0 (42.0-583.0) | 0.688 |
| C-reactive protein, mg/L | 56.0 (0.1-212.0) | 11.1 (0.1-204.5) | <0.001 | 70.8 (0.1-212.0) | 21.0 (0.1-204.5) | 0.078 | 18.1 (9.7-126.0) | 8.1 (0.4-123.0) | <0.05 |
| D-dimer, mg/L | 1.3 (0.2-20.0) | 0.5 (0.1-20.0) | <0.05 | 1.7 (0.2-20.0) | 0.6 (0.1-20.0) | 0.120 | 0.4 (0.2-1.3) | 0.4 (0.2-8.7) | 0.679 |
| Creatine kinase, U/L | 400.0 (203.0-12216.0) | 58.5 (8.8-212.0) | <0.001 | 525.0 (203.0-12216.0) | 64.0 (8.8-212.0) | <0.001 | 231.0 (206.0-1260.0) | 57.5 (19.0-184.0) | <0.001 |
| Lactate dehydrogenase, U/L | 415.0 (147.0-743.0) | 229.0 (2.2-908.0) | <0.001 | 442.0 (147.0-743.0) | 285.0 (2.2-880.0) | <0.01 | 315.0 (210.0-666.0) | 213.0 (2.5-908.0) | <0.05 |
| Alanine aminotransferase, U/L | 44.0 (14.0-173.0) | 25.0 (5.0-1933.0) | <0.01 | 50.0 (14.0-173.0) | 30.0 (5.0-1933.0) | <0.05 | 28.0 (16.0-63.0) | 23.0 (6.0-261.0) | 0.495 |
| Aspartate aminotransferase, U/L | 46.0 (22.0-209.0) | 25.0 (8.0-8191.0) | <0.001 | 53.0 (26.0-209.0) | 31.0 (8.0-8191.0) | <0.001 | 34.5 (22.0-60.0) | 23.0 (9.0-244.0) | 0.065 |
| Blood urea nitrogen, mmol/L | 4.8 (2.0-48.1) | 4.1 (1.5-19.1) | <0.05 | 4.8 (2.4-48.1) | 4.6 (1.5-19.1) | 0.256 | 5.9 (2.0-10.6) | 3.8 (1.6-13.7) | 0.264 |
| Creatinine, µmol/L | 80.5 (43.7-1299.2) | 66.9 (35.9-9435.0) | <0.01 | 81.5 (43.7-1299.2) | 68.4 (35.9-9435.0) | <0.05 | 77.4 (60.7-229.1) | 65.1 (39.4-215.3) | 0.050 |

*P* values indicate differences between patients with and without muscle injury. *P*<0.05 was considered statistically significant.

For the severe subgroup, patients with muscle injury had increased inflammatory response (decreased lymphocyte counts and increased C-reactive protein levels), and more serious liver (increased lactate dehydrogenase, alanine aminotransferase and aspartate aminotransferase levels), kidney (increased creatinine levels) and muscle damage (increased creatinine kinase levels). For non-severe subgroup, patients with muscle injury only had higher C-reactive protein and creatinine kinase levels compared with those without muscle injury.

## Discussion

This is the first report on detailed neurologic manifestations of the hospitalized patients with COVID-19. As of February 19, 2020, of 214 patients included in this study, 88 (41.1%) were severe and 126 (58.9%) were non-severe. Of these, 78 (36.4%) had various neurologic manifestations involved CNS, PNS and skeletal muscles. Compared with non-severe patients, severe patients were older and had more hypertension but less with typical symptoms such as fever and cough. Severe patients were more likely to develop neurological symptoms, especially acute cerebrovascular disease, conscious disturbance and muscle injury. Therefore, for patients with COVID-19, we need to pay close attention to their neurologic manifestations, especially for those with severe infectons, which may have contributed to their demise. Moreover, during the epidemic period of COVID-19, when seeing patients with these neurologic manifestations, doctors should consider SARS-CoV-2 infection as a differential diagnosis so to avoid delayed diagnosis or misdiagnosis and prevention of transmission.

Recently, ACE2 is identified as the functional receptor for SARS-CoV-2 [3], which is present in multiple human organs, including nervous system and skeletal muscle [11]. The expression and distribution of ACE2 remind us that the SARS-CoV-2 may cause some neurological symptoms through direct or indirect mechanisms.

Neurological injury has been confirmed in the infection of other coronavirus such as in SARS-CoV and MERS-CoV. The researchers detected SARS-CoV nucleic acid in the cerebrospinal fluid of those patients and also in their brain tissue on autopsy [12-13].

CNS symptoms were the main form of neurological injury in patients with COVID-19 in this study. The pathological mechanism may be from the CNS invasion of SARS-CoV-2, similar to SARS and MERS virus. Like other respiratory viruses, SARS-COV-2 may enter the CNS through the hematogenous or retrograde neuronal route. The latter can be supported by the fact that some patients in this study had hyposmia. We also found that the lymphocyte counts were lower for patients with CNS symptoms than without CNS symptoms. This phenomenon may be indicative of the immunosuppression in COVID-19 patients with CNS symptoms, especially in the severe subgroup. Moreover, we found severe patients had higher D-dimer levels than that of non-severe patients. This may be the reason why severe patients are more likely to develop cerebrovascular disease.

Consistent with the previous studies [7] muscle symptom was also common in our study. We speculate that the symptom was due to skeletal muscle injury, as confirmed by elevated creatine kinase levels. We found that patients with muscle symptoms had higher creatine kinase and lactate dehydrogenase levels than those without muscle symptoms. Furthermore, creatine kinase and lactate dehydrogenase levels in severe patients were much higher than those of none-severe patients. This injure could be related to ACE2 in skeletal muscle [14]. However, SARS-CoV, using the same receptor, was not detected in skeletal muscle by post-mortem examination [15]. Therefore, whether SARS-CoV-2 infects skeletal muscle cells by binding with ACE2 requires to be further studied. One other reason was the infection-mediated harmful immune response that caused the nervous system abnormalities. Significantly elevated pro-inflammatory cytokines in serum may cause muscle damage.

This study has several limitations. First, only 214 patients were studied, which could cause biases in clinical observation. It would be better to include more patients from Wuhan, other cities in China, and even other countries. Second, all data were abstracted from the electronic medical records, certain patients with neurological problem might not be captured if their neurological symptoms were too mild, such as with hypogeusia and hyposmia. Third, because most patients were still hospitalized and information regarding clinical outcomes was unavailable at the time of analysis, it was difficult to assess the effect of these neurologic manifestations on their outcome, and continued observations of the natural history of disease are needed.

In conclusion, SARS-CoV-2 may infect nervous system, skeletal muscle as well as respiratory tract. In those with severe infection, neurological involvement is more likely, which includes acute cerebrovascular diseases, conscious disturbance and skeletal muscle injury. Involvement of the nervous system carries a poor prognosis. Their clinical conditions may worsen and patients may die soon. Therefore, for patient with COVID19, physicians should pay close attention to any neurologic manifestations in addition to the symptoms of respiratory system.

## Data Availability

All the data referred to in the manuscript are availably.

